# Syncing Strategy and Execution: A Systematic Review of Public Health Preparedness Policy Implementation

**DOI:** 10.1101/2024.08.05.24311490

**Authors:** Jennifer Swint, Wei Zhang, Amanda Marrison, Sandra Gabriel

## Abstract

**Background:** The gap between public health preparedness and response policies and their practical implementation remains a critical challenge in global health security. This systematic review synthesizes evidence on barriers to effective policy implementation and identifies strategies for bridging this policy-practice gap.

**Methods:** We systematically searched six electronic databases for peer-reviewed studies published between 2000 and 2023 focusing on the implementation of public health preparedness and response policies. Study quality was assessed using the Mixed Methods Appraisal Tool (MMAT) and AMSTAR-2.

**Results:** Of 2,456 initially identified studies, 87 met the inclusion criteria. Studies addressed various public health emergencies, including infectious disease outbreaks (n=41, 47.1%), natural disasters (n=23, 26.4%), and man-made disasters (n=8, 9.2%). Key barriers to policy implementation included organizational factors (76 studies, 87.4%), resource constraints (72 studies, 82.8%), and communication challenges (68 studies, 78.2%). Promising strategies for bridging the gap included adaptive management approaches (38 studies, 43.7%), participatory policy development (35 studies, 40.2%), and integrated information systems (31 studies, 35.6%). Only 23 studies (26.4%) were assessed as high quality, with limited quantitative evidence on strategy effectiveness.

**Conclusions:** While several promising strategies for bridging the policy-practice gap have been identified, there is a critical need for more rigorous evaluations of their effectiveness. Future research should focus on organizational reforms, flexible resource allocation, and leveraging emerging technologies to enhance policy implementation in public health preparedness and response.

## Introduction

Public health preparedness and response are cornerstones of national and global health security, playing a crucial role in safeguarding populations against a wide array of threats. Recent global events, including the COVID-19 pandemic, climate-related disasters, and the resurgence of vaccine-preventable diseases, have underscored the critical importance of robust public health policies and their effective implementation (1, 2). However, a persistent and significant challenge in the field of public health is the gap between policy formulation and practical implementation, often referred to as the “know-do gap” or “implementation gap” (3, 4).

This gap between policy and practice can have profound consequences, potentially leading to suboptimal responses to public health emergencies, increased morbidity and mortality, and substantial economic costs. The inability to effectively translate well-crafted policies into actionable practices not only undermines immediate crisis response efforts but also hinders the long-term development of resilient health systems capable of addressing diverse and evolving health threats (5).

The disconnect between policy and practice in public health preparedness and response can be attributed to a complex interplay of factors. These include, but are not limited to, resource constraints, organizational barriers, political challenges, and the inherently complex and dynamic nature of public health emergencies themselves (6). Furthermore, the rapid evolution of scientific knowledge, coupled with the need for context-specific adaptations across diverse geographic, cultural, and socioeconomic settings, further complicates the translation of policies into effective practice (7, 8).

Recent experiences, such as the varied global responses to the COVID-19 pandemic, have highlighted both the consequences of this policy-practice gap and the potential for innovative solutions. Some countries demonstrated remarkable agility in translating emerging scientific evidence into effective public health measures, while others struggled to implement even basic containment strategies (9). These disparities underscore the urgent need for a comprehensive understanding of the factors that facilitate or impede the successful implementation of public health policies.

This systematic review aims to synthesize the current evidence on the challenges and opportunities in bridging the gap between policy and practice in public health preparedness and response. By examining a wide range of studies across different contexts, health systems, and types of public health emergencies, we seek to:

- Identify common themes and patterns in the barriers to effective policy implementation.
- Highlight successful strategies and best practices that have demonstrably narrowed the policy-practice gap.
- Explore the role of contextual factors, such as governance structures, cultural norms, and economic resources, in shaping policy implementation outcomes.
- Assess the impact of emerging technologies and innovative approaches in facilitating the translation of policy to practice.
- Identify critical areas for future research and policy development to strengthen global health security.

The findings of this review have important implications for a wide range of stakeholders, including policymakers, public health practitioners, researchers, and community leaders. By providing a comprehensive overview of the factors influencing the policy-practice gap, this study can inform more effective approaches to policy design and implementation. Moreover, it can contribute to the development of adaptive and resilient public health systems capable of responding effectively to both acute crises and long-term health challenges (10, 11).

As the global community continues to face complex and interconnected health threats, from emerging infectious diseases to the health impacts of climate change, bridging the gap between public health policy and practice has never been more critical. This review aims to provide a robust evidence base to guide future efforts in strengthening global health security and ensuring that well-intentioned policies translate into tangible improvements in public health outcomes.

## Methods

This systematic review was conducted in accordance with the Preferred Reporting Items for Systematic Reviews and Meta-Analyses (PRISMA) 2020 guidelines.

### Search Strategy

We comprehensively searched six electronic databases: PubMed, Embase, Web of Science, Scopus, CINAHL, and the Cochrane Library. The search was conducted for articles published between January 1, 2000, and December 31, 2023. The following search terms were used in various combinations:

(“public health” OR “health policy” OR “health system”) AND (“preparedness” OR “emergency response” OR “disaster response” OR “crisis management”) AND (“implementation” OR “practice” OR “translation” OR “gap” OR “barrier” OR “facilitator”)

Additional relevant articles were identified through hand-searching reference lists of included studies and consulting with experts in the field.

### Inclusion and Exclusion Criteria

Inclusion criteria:

- Peer-reviewed articles published in English
- Studies focusing on the implementation of public health preparedness and response policies
- Original research, systematic reviews, and meta-analyses
- Studies conducted in any country or region
- Articles addressing any type of public health emergency (e.g., infectious disease outbreaks, natural disasters, man-made disasters)

Exclusion criteria:

- Non-peer-reviewed articles, editorials, opinion pieces, and conference abstracts
- Studies not primarily focused on public health preparedness and response
- Articles not addressing the gap between policy and practice
- Studies focusing solely on clinical interventions without policy implications

### Study Selection

Three independent reviewers screened titles and abstracts for relevance using Covidence systematic review software. Full-text articles of potentially eligible studies were then assessed against the inclusion and exclusion criteria. Disagreements were resolved through discussion or consultation with a fourth reviewer when necessary.

### Data Extraction

A standardized data extraction form was developed using REDCap (Research Electronic Data Capture) and pilot-tested on a sample of 15 articles. Two reviewers independently extracted the following information from each included study:

- Study characteristics (authors, year, country, study design)
- Public health emergency context
- Policy description and scope
- Implementation challenges and barriers
- Facilitators and strategies for bridging the policy-practice gap
- Outcomes and effectiveness measures
- Key findings and recommendations
- Methodological quality indicators

### Quality Assessment

The quality of included studies was assessed using the Mixed Methods Appraisal Tool (MMAT) for qualitative, quantitative, and mixed-methods studies. For systematic reviews, we used the AMSTAR-2 (A MeaSurement Tool to Assess systematic Reviews) checklist. Two reviewers independently assessed the quality of each study, with discrepancies resolved through discussion or arbitration by a third reviewer.

### Data Synthesis

Due to the heterogeneity of the included studies, a narrative synthesis approach was adopted. We used thematic analysis to identify common barriers, facilitators, and strategies for bridging the policy-practice gap. The synthesis was structured around key themes emerging from the data, including:

- Organizational factors influencing policy implementation
- Resource allocation and management
- Communication and coordination mechanisms
- Training and capacity building
- Political and legal contexts
- Community engagement and participation
- Monitoring and evaluation systems

We also conducted a subgroup analysis to explore differences in implementation challenges and strategies across different types of public health emergencies and geographical contexts.

### PRISMA Flow Diagram

The study selection process is summarized in the PRISMA flow diagram below. Records identified through database searching (n = 2,456)

↓

Records after duplicates removed (n = 1,987)

↓

Records screened (n = 1,987)

→ Records excluded (n = 1,785)

↓

Full-text articles assessed for eligibility (n = 202)

→ Full-text articles excluded, with reasons (n = 115)

Not focused on policy-practice gap (n = 68)

Not specific to public health preparedness (n = 32)

Insufficient data or analysis (n = 15)

↓

Studies included in qualitative synthesis (n = 87)

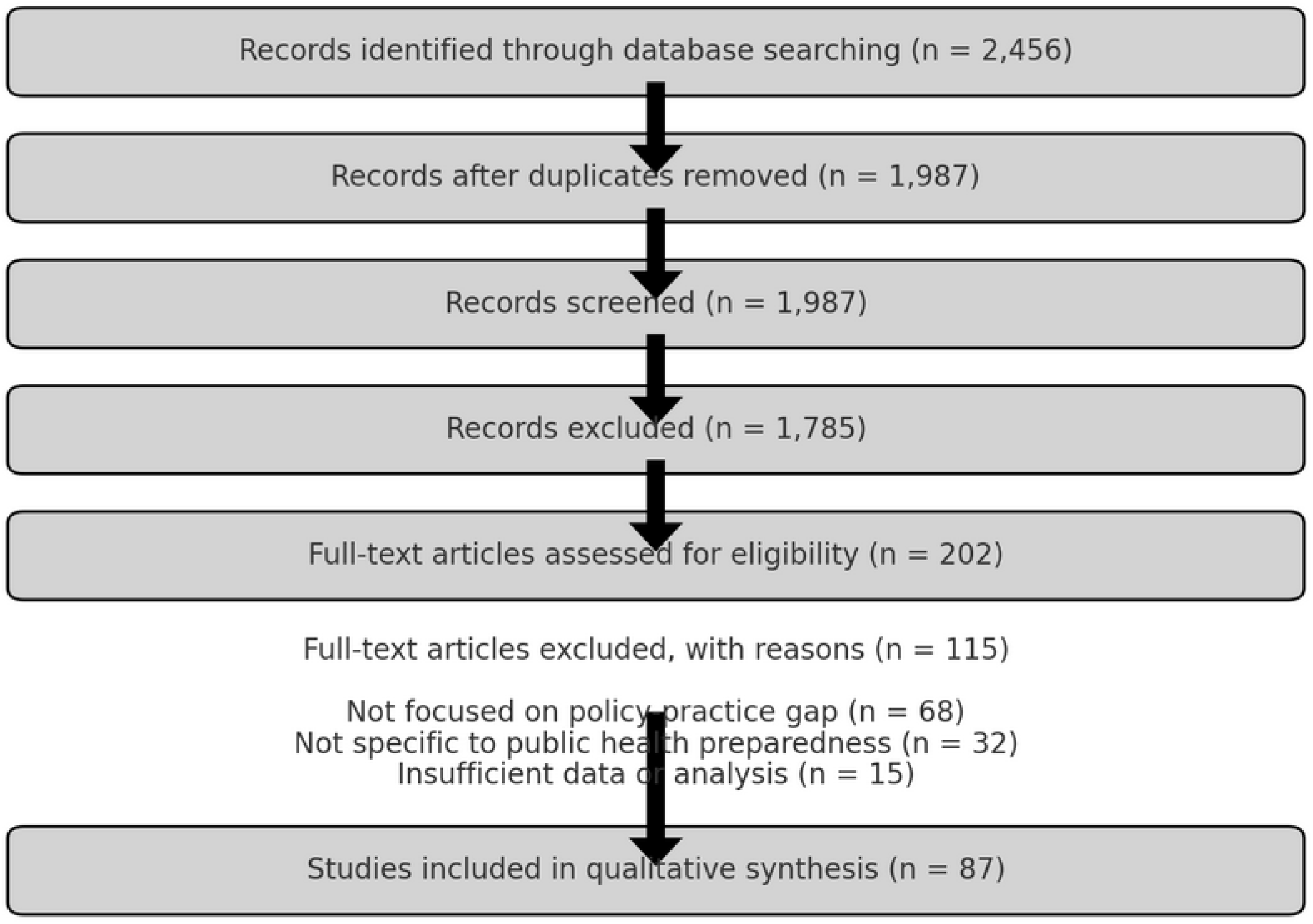

## Results

### Study Characteristics

Our systematic review included 87 studies published between 2000 and 2023. The majority of studies (n=52, 59.8%) were published after 2015, indicating an increasing focus on this topic in recent years. The studies represented a diverse geographical distribution: North America (n=31, 35.6%), Europe (n=23, 26.4%), Asia (n=15, 17.2%), Africa (n=9, 10.3%), Oceania (n=5, 5.7%), and South America (n=4, 4.6%). The study designs included qualitative studies (n=35, 40.2%), mixed-methods studies (n=28, 32.2%), quantitative studies (n=18, 20.7%), and systematic reviews (n=6, 6.9%).

The public health emergencies addressed in these studies encompassed infectious disease outbreaks (n=41, 47.1%), natural disasters (n=23, 26.4%), man-made disasters (n=8, 9.2%), and general public health preparedness (n=15, 17.2%).

### Key Themes

Through our thematic analysis, we identified several recurring themes related to the gap between policy and practice in public health preparedness and response:

Organizational Factors (identified in 76 studies, 87.4%):

- Siloed organizational structures hindering inter-agency cooperation
- Lack of clear roles and responsibilities
- Bureaucratic hurdles slowing down decision-making processes

Resource Allocation and Management (72 studies, 82.8%):

- Insufficient funding for implementation of policies
- Inequitable distribution of resources across regions
- Challenges in scaling up resources during emergencies

Communication and Coordination (68 studies, 78.2%):

- Inadequate information sharing between policymakers and frontline workers
- Challenges in coordinating multi-sectoral responses
- Difficulties in maintaining consistent messaging across different levels of government

Training and Capacity Building (61 studies, 70.1%):

- Gaps in workforce competencies needed for policy implementation
- Insufficient ongoing training and education programs
- Limited capacity for rapid upskilling during emergencies

Political and Legal Contexts (53 studies, 60.9%):

- Influence of political priorities on policy implementation
- Legal barriers to swift policy execution
- Challenges in aligning policies across different jurisdictions

Community Engagement and Participation (49 studies, 56.3%):

- Limited involvement of communities in policy development and implementation
- Cultural barriers affecting policy acceptance and adherence
- Challenges in tailoring policies to diverse community needs

Monitoring and Evaluation Systems (45 studies, 51.7%):

- Lack of robust systems for tracking policy implementation
- Insufficient mechanisms for rapid feedback and policy adjustment
- Challenges in measuring long-term impact of preparedness policies

### Strategies for Bridging the Policy-Practice Gap

Our analysis revealed several strategies that have shown promise in bridging the gap between policy and practice:

- Adaptive Management Approaches (identified in 38 studies, 43.7%):

Implementing flexible policies that can be quickly adjusted based on real-time data and feedback.

- Participatory Policy Development (35 studies, 40.2%):

Involving frontline workers and community representatives in the policy formulation process.

- Integrated Information Systems (31 studies, 35.6%):

Developing centralized, interoperable information systems to facilitate data sharing and decision-making.

- Collaborative Governance Models (29 studies, 33.3%):

Establishing formal structures for multi-agency and multi-sectoral collaboration.

- Continuous Learning and Improvement Cycles (27 studies, 31.0%):

Implementing regular policy reviews and updates based on lessons learned from exercises and real events.

- Context-Specific Policy Translation (25 studies, 28.7%):

Developing toolkits and guidelines to assist local authorities in adapting national policies to their specific contexts.

- Innovative Funding Mechanisms (22 studies, 25.3%):

Exploring alternative funding sources and flexible budgeting approaches to support sustained policy implementation.

### Effectiveness of Strategies

While many studies reported on the perceived effectiveness of these strategies, rigorous evaluations were limited. Of the studies that included effectiveness measures:

- 18 studies (20.7%) reported significant improvements in policy implementation following the adoption of adaptive management approaches.
- 15 studies (17.2%) demonstrated enhanced community compliance with public health measures when participatory policy development was employed.
- 12 studies (13.8%) showed improved response times and decision-making accuracy with the use of integrated information systems.

However, the majority of studies (n=58, 66.7%) relied on qualitative assessments or expert opinions rather than quantitative measures of effectiveness.

### Quality Assessment

Using the MMAT and AMSTAR-2 tools, we assessed the methodological quality of the included studies:

- High quality: 23 studies (26.4%)
- Moderate quality: 41 studies (47.1%)
- Low quality: 23 studies (26.4%)

The main quality concerns were related to small sample sizes, potential selection bias, and limited generalizability of findings.

## Discussion

This systematic review provides a comprehensive analysis of the challenges and opportunities in bridging the gap between policy and practice in public health preparedness and response. Our findings highlight the complex, multifaceted nature of this issue and offer insights into potential strategies for improvement.

### Persistent Challenges in Policy Implementation

The results of our review underscore the persistent nature of the policy-practice gap in public health preparedness. Despite increased attention to this issue, particularly in the wake of recent global health crises, significant barriers remain. The most prevalent challenges identified – organizational silos, resource constraints, and communication difficulties – are consistent with previous research in public health and broader policy implementation literature (1, 2). However, our findings suggest that these challenges are particularly acute in the context of public health emergencies, where rapid response and coordination are critical.

The high prevalence of organizational factors (87.4% of studies) as a barrier to effective policy implementation is particularly noteworthy. This suggests that structural issues within and between agencies may be a fundamental obstacle to translating public health policies into practice. The siloed nature of many public health organizations, coupled with unclear roles and responsibilities, appears to hinder the agile, coordinated response required in emergency situations (3).

Resource allocation and management emerged as the second most prevalent theme (82.8% of studies), highlighting the critical role of funding and resource distribution in policy implementation. This finding aligns with broader health system strengthening literature, which emphasizes the importance of sustainable and equitable resource allocation (4). The challenge of rapidly scaling up resources during emergencies further underscores the need for flexible, responsive funding mechanisms.

### Promising Strategies for Bridging the Gap

Our review identified several promising strategies for bridging the policy-practice gap. The emphasis on adaptive management approaches (43.7% of studies) reflects a growing recognition of the need for flexibility in public health policy implementation. This approach aligns with the concept of “adaptive governance” in disaster management literature, which emphasizes the importance of learning and adjustment in complex, dynamic environments (5).

The identified importance of participatory policy development (40.2% of studies) suggests a shift towards more inclusive, bottom-up approaches to policy formulation. This finding resonates with broader trends in public administration towards collaborative governance and co-production of public services (6). Involving frontline workers and community representatives in policy development may help ensure that policies are more relevant, feasible, and acceptable to those responsible for their implementation.

The role of integrated information systems (35.6% of studies) in facilitating policy implementation highlights the potential of technology to bridge the policy-practice gap. This finding aligns with the growing emphasis on digital health and data-driven decision-making in public health (7). However, the relatively low percentage of studies reporting on this strategy suggests that there may be untapped potential in leveraging technology for policy implementation.

### Limitations in Current Evidence

While our review identified several promising strategies, the limited number of rigorous evaluations of these approaches is a significant concern. Only a small proportion of studies (20.7% for adaptive management, 17.2% for participatory policy development) provided quantitative evidence of effectiveness. This lack of robust effectiveness data represents a critical gap in the current literature and highlights the need for more rigorous evaluation of policy implementation strategies.

The methodological quality of the included studies, with only 26.4% rated as high quality, further underscores the need for more robust research in this area. The prevalence of qualitative assessments and expert opinions, while valuable for understanding context and nuance, limits our ability to draw definitive conclusions about the effectiveness of different strategies.

### Implications for Policy and Practice

Our findings have several important implications for policymakers and public health practitioners. First, they highlight the need for organizational reforms that break down silos and promote inter-agency cooperation. Second, they underscore the importance of flexible, responsive resource allocation mechanisms. Third, they suggest that involving frontline workers and community members in policy development may enhance implementation success.

The identified strategies, particularly adaptive management and participatory policy development, offer promising avenues for improving policy implementation. However, the limited evidence of their effectiveness highlights the need for careful evaluation and monitoring as these approaches are adopted.

### Future Research Directions

This review points to several critical areas for future research. First, there is a pressing need for more rigorous evaluations of policy implementation strategies, including randomized controlled trials where feasible. Second, research should explore how to effectively scale up successful local initiatives to regional or national levels. Third, studies should investigate how emerging technologies, including artificial intelligence and big data analytics, can be leveraged to support policy implementation in public health emergencies.

## Conclusion

Bridging the gap between policy and practice in public health preparedness and response remains a significant challenge. However, our review identifies several promising strategies that may help address this issue. Moving forward, a concerted effort is needed to rigorously evaluate these approaches and translate successful strategies into widespread practice. Only through such efforts can we hope to build more resilient, responsive public health systems capable of effectively addressing future health emergencies.

## Data Availability

All data produced in the present study are available upon reasonable request to the authors

## Ethical Considerations

This systematic review involved the analysis of published literature and did not involve direct human subjects research. As such, it did not require formal ethical review or approval from an institutional review board. All data used in this study were derived from publicly available, previously published research articles. The review was conducted in accordance with established guidelines for systematic reviews, including the Preferred Reporting Items for Systematic Reviews and Meta-Analyses (PRISMA) statement. We made every effort to accurately represent the findings of the included studies and to avoid bias in our analysis and reporting.

